# Neural Fragility of the Intracranial EEG Network Decreases after Surgical Resection of the Epileptogenic Zone

**DOI:** 10.1101/2021.07.07.21259385

**Authors:** Adam Li, Patrick Myers, Nebras Warsi, Kristin M. Gunnarsdottir, Sarah Kim, Viktor Jirsa, Ayako Ochi, Hiroshi Otusbo, George M. Ibrahim, Sridevi V. Sarma

## Abstract

Over 15 million patients with epilepsy worldwide do not respond to medical therapy and may benefit from surgical treatment. In focal epilepsy, surgical treatment requires complete removal or disconnection of the epileptogenic zone (EZ). However, despite detailed multimodal pre-operative assessment, surgical success rates vary and may be as low as 30% in the most challenging cases. Here we demonstrate that neural fragility, a dynamical networked-system biomarker of epileptogenicity, decreases following successful surgical resection. Moreover, neural fragility increases or remains constant when seizure-freedom is not achieved. We demonstrate this retrospectively in a virtual patient with epilepsy using the Virtual Brain neuroinformatics platform, and subsequently on six children with epilepsy with pre- and post-resection intra-operative recordings. Finally, we compare neural fragility as a putative biomarker of epileptogenicity against established spectral metrics, such as high frequency oscillations and find that neural fragility is a superior biomarker of epileptogenicity.

## 1 Introduction

Drug-resistant epilepsy (DRE), where seizures continue despite adequate trial of at least two tolerated appropriately chosen anti-epileptic drugs, affects over 15M patients worldwide and 1M in the US.^1^ Patients with DRE have an increased risk of sudden are burdened by epilepsy-related disabilities, and the cost of their care and frequent hospitalizations is a significant contributor to the $16 billion spent annually in the US treating epilepsy patients.^2^ Approximately 50% of patients with DRE have focal epilepsy and may be amenable to surgical treatment via resection or disconnection of the epileptogenic zone EZ), which typically involves the seizure onset zone (SOZ) and early propagation zone (EPZ).

Localizing the EZ is an ill-posed task as it is impossible to identify despite multimodal preoperative assessments. The common definition requires one to resect the hypothesized epileptogenic brain tissue and then verify that a patient is seizure-free to be certain that the EZ was correctly identified and was within the subset of resected tissue.^3^ In cases of seizure-freedom, it is impossible to be certain whether a smaller resection may have achieved similar outcomes. These outcomes are typically measured at least 6-12 months from surgery based on the Engel score, or ILAE classification.^4,5^

If seizures reoccur, re-operations have increased risk of permanent postoperative neurological deficits,^6^ increased health-care costs (32% higher than in patients with successful first-time surgeries^7^), and subsequent surgeries may yield a lower likelihood of success.^8^ When localizing the EZ, the focus is on identifying the SOZ and EPZ from scalp and intracranial EEG (iEEG) recordings captured during seizures. As a result, there are lengthy hospital stays and increased risks of complications.^9,10^ Interictal localization using iEEG typically relies on visual identification of “epileptic signatures”, such as beta-buzz, high-frequency oscillations (HFOs), or interictal spiking.^11,12,13^ These often yield an estimate of the irritative zone, which does not always correlate to the SOZ. Due to imprecise EZ localization, healthy neural tissue may be included within resection margins potentially resulting in avoidable neurological deficit.^14,15,16^ Furthermore, surgical resection may yield poor outcomes in the most challenging cases with success rates varying between 30-70%.^17^

Several EZ localization algorithms have been proposed to better leverage iEEG. Many entail investigations of the spectral power in each iEEG channel, including HFOs.^18^ However, these approaches do not consider network properties of the brain because they treat each EEG channel independently. Others have proposed graph-based analysis of iEEG,^19,20,21,22,23^ but these approaches fail to identify internal network properties that cause seizures to occur in the first place. Moreover, underlying graph and connectivity structure is typically not observed, but estimated from data. These methods define a dependency measures such as Pearson correlation,^22^ or coherence.^24^ However, any noise in the estimation procedure can drastically alter the underlying graph metrics, such as centrality, or degree. In,^25^ it is shown that that graph measures are not unique to a graph. Many graphs have the same degree distribution but are completely different graphs.

Neural fragility, a structured perturbation of a networked dynamical system estimated from ictal iEEG data has the potential to be a robust biomarker of the EZ.^26,27,28^ It was validated in 91 patients across five clinical centers and outperformed the most popular channel and staticnetwork-based features. This model is dynamic, based on robust least-squares estimation^29^ and models intrinsic stability properties of the system. That is, the propensity for seizures in our model occur as a property of eigenvalue perturbations. However, there is still an unmet need to develop more robust interictal biomarkers of the EZ to determine the extent of a resection required in the OR, thereby optimizing the surgical outcome, while limiting the amount of brain tissue removed. The presence of a robust interictal biomarker could also facilitate and expand the utility of intraoperative electrocorticography. At present, interictal analysis entails the detection of interictal discharges, with limited utility. A biomarker that leverages interictal data and specific to the EZ would significantly advance surgical approaches for focal epilepsy.^30,31^ In this study, we evaluate neural fragility as a correlate of surgical outcome using pre and post resection iEEG interictal recordings of the same subjects. We hypothesize that neural fragility of the brain network will decrease in cases of successful surgical resection. In iEEG data, neural fragility will modulate with respect to surgical outcome, increasing or staying the same after a failed resection (i.e., residual epileptic tissue shows fragility), and decreasing after resection if the procedure was successful. We first test this hypothesis by using the Virtual Brain (TVB) simulation platform and a virtual epileptic patient model to simulate various scenarios of failed and successful resections, and compute neural fragility of the simulated pre/post resection recordings. We demonstrate that neural fragility decreases overall in the iEEG network when the entire EZ is removed, partially decreases when some of the EZ is removed, and increases or remains unchanged when none of the EZ is removed. We further present data supporting this hypothesis with a dataset of 6 patients (5 success and 1 failure outcome) selected from the Hospital for Sick Children (HSC). Since we are analyzing interictal data, we also compute HFOs using a root-mean square (RMS) detector. We demonstrate that neural fragility predicts outcomes better compared to more established frequency band powers, or HFOs. Our results suggest that neural fragility may be helpful in predicting surgical outcome and the extent of the resected area using pre and post-resection iEEG recordings. This can potentially improve surgical success rates and allow surgeons to incrementally operate on the EZ thereby limiting the amount of brain tissue needed to be resected for the patient to be seizure free.

## 2 Materials and Methods

### 2.1 Ethics Statement

Decisions regarding the need for invasive monitoring and the placement of electrode arrays were made independently of this work and part of routine clinical care. All data were acquired with ethics approval from the Research Ethics Board (REB) at the Hospital of Sick Children. The acquisition of data for research purposes was completed using guidelines established in accordance with the Code of Ethics of the World Medical Association (1964, Declaration of Helsinki).

We confirm that we have read the Journal’s position on issues involved in ethical publication and affirm that this report is consistent with those guidelines.

### 2.2 The Hospital for Sick Children

iEEG data from 6 DRE patients were analyzed. These patients underwent intracranial EEG monitoring between January 2017 and December 2019 at The Hospital for Sick Kids (HSC). All children underwent subdural grid implantation and the same chronically implanted electrodes were used without changing position or orientation for extraoperative and subsequent intraoperative electrocorticography, using a previously reported technique.^32^ For full details of the dataset, see Supplementary Materials and Methods.

### 2.3 The Virtual Brain Data

We also used a single virtual epileptic patient for our simulation analysis with The Virtual Brain. One patient from^33^ were used. Neuroimaging data, and specifically diffusion MRI were collected for this subject and a full connectivity dataset was constructed that would allow TVB simulations. All acquisition information for this subject can be found in.^33^ We list the patient metadata used in this paper in the Supplementary table. Details on the imaging and implantation data to instantiate the virtual epileptic patient are presented in Supplementary Materials.

### 2.4 Simulating a Virtual Epileptic Patient

The Virtual Brain (TVB) is a neuroinformatics platform used to simulate whole-brain neural dynamics. It incorporates biologically realistic computational models and simulates brain network dynamics using connectome-based approaches and directly linking them to various brain imaging modalities.^34^ We use the resting-state Epileptor model, designed to simulate resting state interictal activity. We set a region to be EZ, with the rest normal and then measure the iEEG output of the TVB model. This data consists of the pre-resection simulation. Afterwards, we simulate data from three resective scenarios that fully captures the EZ, partially captures the EZ, or completely fails to capture the EZ. For full details of the simulation, see Supplementary Material.

### 2.5 Neural Fragility Analysis

Neural fragility is a concept based on the conjecture that focal seizures arise from a few fragile nodes, i.e., the, which renders the cortical epileptic network on the brink of instability.^35^ When one observes iEEG data during interictal, or preictal periods, activity recorded from each channel *appears* to hover around a baseline value. If the network is “balanced”, then it will respond transiently to a perturbation or stimulus, but always returns back to the baseline value. In contrast, when one observes iEEG data during a seizure event, activity (i) grows in amplitude, (ii) oscillates, and (iii) spreads in the brain. We model this susceptibility of a seizure event within the iEEG network.

Our conjecture is that small changes in connection strengths at an EZ node causes an imbalance in inhibitory and excitatory connectivity between brain regions and is thus “fragile”. Either inhibition is decreased and/or excitation is increased due to a change in connection strengths between the EZ node and its neighbors. We compute fragility heatmaps for every single patient and simulated-patient dataset. We compared fragility against time-frequency analysis of the data. For full details of analysis, see Supplementary Materials.

## 3 Results

In this work, we analyze pre and post resection iEEG data and compute neural fragility of the iEEG network to compare the changes in fragility stratified by the actual surgical outcome (see Figure 1). iEEG provides high temporal resolution data that enables detection of abnormal activity, such as spikes and high frequency bursts, in between seizures (interictal) and during seizures (ictal). Moreover, intraoperative data may refine and modify the resection plan in real-time, by capturing interictal data to study electrophysiological changes within the irritative zone following resection. iEEG provides high temporal resolution data that enables clinicians to visually detect abnormal activity, such as spikes and high frequency bursts, in between seizures (interictal) and during seizures (ictal). Moreover, intraoperative data may refine and modify the resection plan in real-time, by capturing primarily interictal data to study electrophysiological changes within the irritative zone following resection.^32,10^ To extend our work in,^26,27^ we hypothesize that neural fragility of iEEG data will modulate with respect to the successful surgical resection of the EZ.

**Figure 1.**
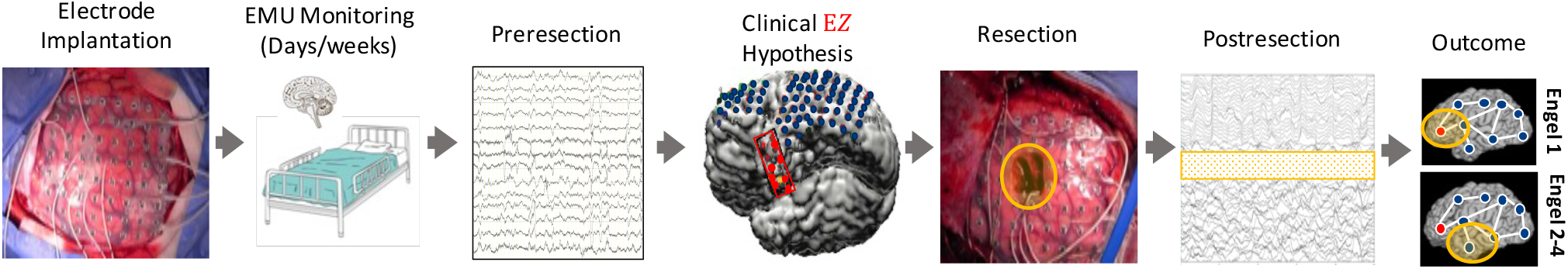
Clinical workflow with continuous iEEG monitoring before and after a surgical resection. An overview of the DRE treatment clinical procedure. Patients are accepted into the Epilepsy Monitoring Unit (EMU) and implanted with intracranial electrodes to undergo monitoring. They are typically in the EMU for many days, up to a few weeks. Pre-resection iEEG data is used to form a clinical EZ hypothesis (red circled region). The clinical EZ is an estimate of the true EZ, and may contain the EZ, or not at all. Based on the clinical EZ hypothesis, a surgical resection is subsequently performed to remove that region of the brain (orange circled region). This is evaluated post-hoc (i.e. after the surgery is completed), which is why the EZ is difficult to define. There is not clinical biomarker that can define the EZ prospectively, thus the patient outcome after resections determine if clinicians successfully localized the EZ. Immediately afterwards in these patient recordings, post-resection iEEG data is recorded. Patients have followups 12+ months later to determine the actual outcome of the surgical treatment and whether the true EZ was successfully removed. The outcome of the patient is then measured in terms of Engel scores, where I is seizure free, and II-IV represent increasing levels of post-op seizure severity.

To test this hypothesis, we use TVB to generate simulated iEEG data from real patient diffusion tensor imaging (DTI) connectomes, and show how neural fragility modulates with proposed complete, partial, and incomplete resections of the EZ. We then demonstrate that neural fragility modulates pre and post resection in DRE patients that underwent surgical resection at HSC. A neurosurgical procedure at HSC collects intraoperative iEEG using the same chronically implanted electrodes used for extraoperative mapping. These data facilitate resective epilepsy surgery by allowing one to observe the iEEG network while a surgery takes place. Intraoperative electrocorticography (ECoG) monitoring in Warsi, et al.^32^ allows real-time monitoring of the brain and allows for post-resection recordings that from electrodes sampling the identical brain regions. Thus, clinically, one could monitor the patient for a brief period of time after resective surgery to determine if there are any recurring seizures, or epileptic activity.^10^ If one has a proposed marker for the underlying EZ, the model can be validated continuously throughout the operation. This provides a distinct advantage over the classical “resect and wait” retrospective datasets because we have access to post-resection recordings.^32,10^

Finally, we analyze every HSC patient’s iEEG using 7 other frequency-based benchmark features, resulting in spatiotemporal heatmaps for every feature. The baseline features include spectral power in various frequency bands (e.g. delta band 1-4 Hz) and HFOs computed via an root mean-square (RMS) detector. We consider all of these as potential EEG representations of the epileptic network to see if they correlate with surgical outcome based on pre and post resection data.

### 3.1 Neural fragility of an iEEG network

Neural fragility is a concept based on the conjecture that focal seizures arise from a few fragile nodes, i.e., the EZ, which renders the epileptic network on the brink of instability. In,^27^neural fragility is introduced in the context of “balanced” and “imbalanced” networks. Balanced networks respond transiently to an impulse, returning to baseline values, whereas imbalanced networks respond to an impulse with electrical activity that grows in amplitude, oscillates and spreads in the brain. From a dynamical systems perspective, the epileptic iEEG network is an unstable network (capable of seizing). Neural fragility of a node in the epileptic network is defined as the minimum amount of perturbation on the network structure required to move the system from a stable to unstable state. This specific perturbation can take on a variety of forms. In Li et al.,^27^ a column perturbation (modifying the outgoing connections of one node) was applied for EZ localization. In this work, we further this notion and use a row perturbation (modifying the incoming connections of one node) and column perturbation to specify that a region in the epileptic network is fragile (see Figure 2). We compute product-fragility (i.e. the product of the row and column perturbation norms) to determine which nodes are fragile in both a column and row aspect. We hypothesize that epileptic regions are more detectable if they have both high row and column fragility. Because both perturbations are normalized to have norm less than one, taking the product will result in more stable maps. Taking a sliding window over the iEEG data, we estimate a linear dynamical system using least-squares^29^ (see Supplementary Neural Fragility Analysis). Then we compute this over a sliding window of the iEEG data to get a spatiotemporal fragility heatmap.

**Figure 2.**
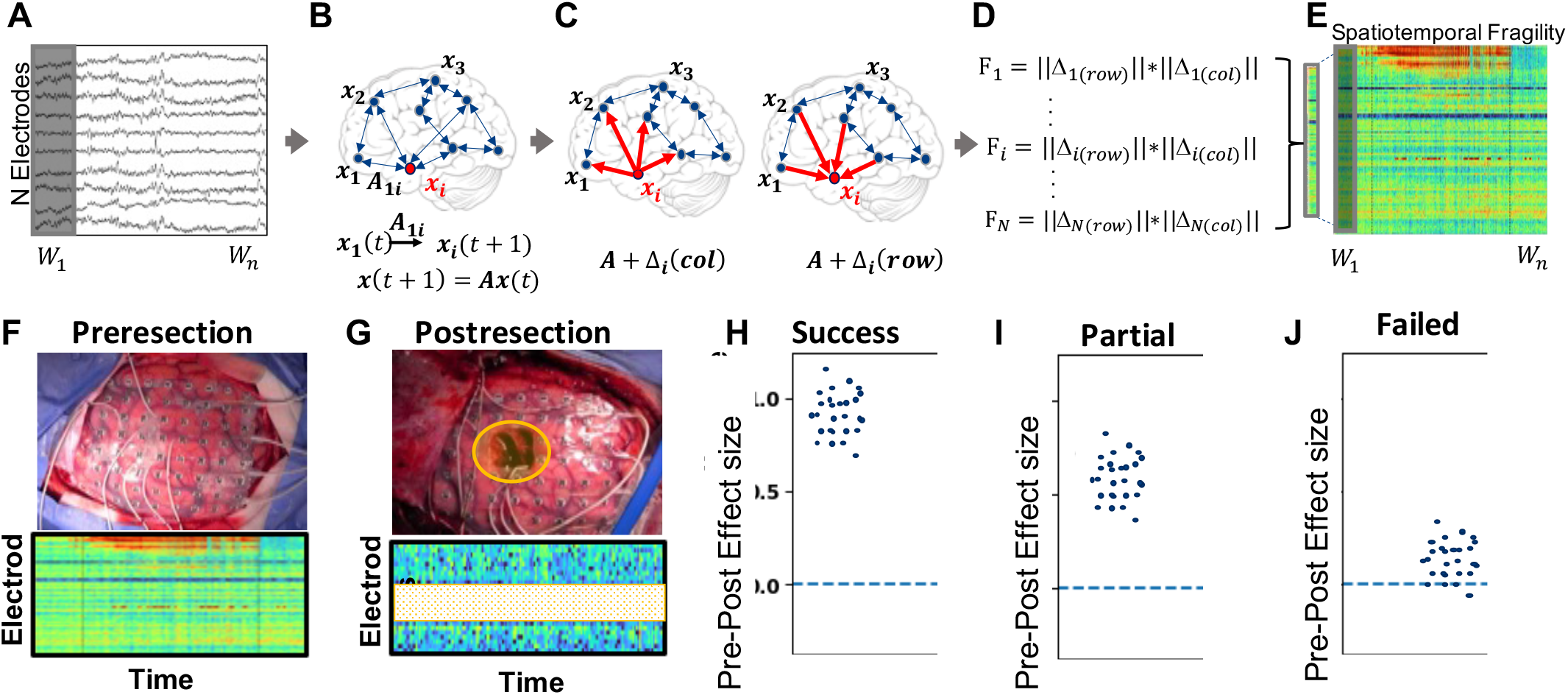
Computing fragility as the product of column and row perturbations. **(A)** From a small time window of N iEEG electrodes, **(B)** a linear time-invariant dynamical system, represented as an A matrix, is estimated. **(C)** Neural fragility is computed as the minimum amount of energy (measured in norm), represented as a Δ matrix, required to destabilize the linear system. This can be computed for every node within the N-node network (i.e. iEEG electrodes). The norm of the Δ matrix can be computed as a column perturbation over the N nodes, where the perturbation matrix computed has a rank-1 structure with 0’s in every column except for the node being perturbed. Similarly, the norm of the Δ matrix can be computed as a row perturbation over the N nodes, where the perturbation matrix computed has a rank-1 structure with 0’s in every row except for the node being perturbed. **(D)** The row and column fragility are combined as a product for all electrodes at every single time point. **(E)** This is then summarized as a spatiotemporal heatmap. **(F)** Taking iEEG data from preresection sessions, we compute heatmaps and then compare these with **(G)** postresection sessions. When we compare the spatiotemporal values between the two sessions using a bootstrap sampling procedure, we expect **(H)** successful surgeries to have a positive effect size. **(I)** Partially failed surgeries, where the EZ is not fully captured, should result in smaller, but still positive effect size. **(J)** Finally, a failed surgery, where the EZ is not resected at all would result in a 0 effect size, or even possibly negative effect size difference between pre and post resection sessions. If a biomarker can detect the presence of the EZ in the network, then one expects it to modulate depending on if the EZ is successfully removed.

### 3.2 Neural fragility modulates after complete, partial and incomplete resection of the EZ in simulation

To determine how neural fragility of the entire observed iEEG network can potentially be used as an estimator for surgical outcome, we first study how the entire networks’ neural fragility modulates in an in-silico environment using TVB. Using the resting-state Epileptor model, we simulate iEEG data using real patient connectomes derived from DTI and T1 MRI data.^34^ We set the EZ regions for each simulation based on the actual clinically hypothesized EZ. For full details on simulations, see The Virtual Brain Patient-Specific Modeling. We model resections by removing that part of the structural connectome. Complete, partial and incomplete resections are modeled as a complete removal of the EZ brain region, partial removal of the EZ, and then a completely incorrect resection of a non-epileptic brain region. For a breakdown of some of the clinical characteristics present in the TVB dataset, see Supplementary Table S2.

In Figure 3a-c, we show the neural fragility heatmap for an example of a complete, partial and incomplete in-silico resection of the EZ. Neural fragility decreases significantly (K-sample MANOVA PValue = 4.16e-7) in the successful resections with a Cohen’s D effect size 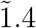 and 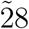 times greater than the partial and incomplete resections respectively. The successful resection resulted in an effect size difference between the overall network fragility of 0.761 ± 0.322, while a failed resection resulted in an effect size of 0.025 ± 0.244.

**Figure 3.**
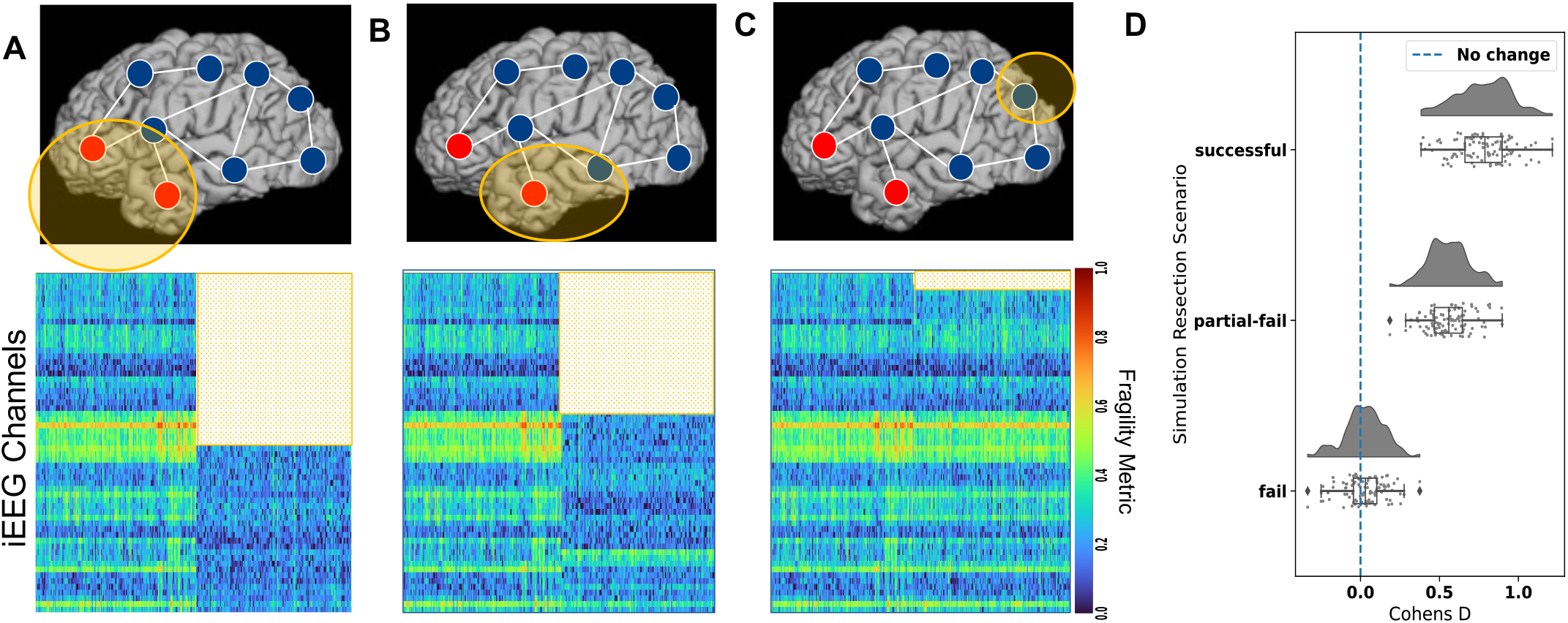
Neural product fragility of complete, partial and incomplete in-silico resections of the EZ (A) Neural fragility heatmap of a successful resection of the underlying EZ. The heatmap shows two concatenated sessions: the pre-resection iEEG and post-resection iEEG. The white region represents the channels that were in the resected regions for the postresection iEEG simulation. **(B)** Neural fragility of a partially successful resection, where one epileptic region was resected, but another one was left in. Values in the post-resection period still go down, but relative to panel (a), they are slightly higher. **(C)** Neural fragility of a completely failed resection, where an incorrect brain region was removed. There is qualitatively very little difference with respect to the pre-resection session. The turbo colormap is used in these heatmaps a-c. **(D)** A summary effect size difference between pre and post resection fragility values for the three resective scenarios from a-c. Each dot represents the Cohens D effect size computed on a bootstrap sample from pre and post resection heatmap. The successful resections have an improvement in overall network fragility (positive Cohen’s D), while the failed resection shows essentially no effect difference. The Cohen’s D effect size of successful, partial, and incomplete resections were 0.761 ± 0.322 (PValue of 4.16e-7), 0.542 ± 0.272 (PValue of 2.19e-5) and 0.025 ± 0.244 (PValue of 4.12e-3) respectively (all effect sizes are 95% confidence interval). All PValues were computed using a K-Sample MANOVA test using distance correlation with 0.05 alpha level. For more information on how the bootstrap procedure was implemented, see Supplementary Statistical Analysis.

### 3.3 Neural fragility decreases in patients with successful resections

Next, we validated our results from TVB simulations on iEEG data from pediatric cases (n=6) with DRE that had epilepsy monitoring and subsequent resective surgery from HSC. This dataset is unique because the chronically same electrodes continue to record throughout the resection, while sampling the identical brain regions.^32^ For a full clinical description of the patients from HSC, see Supplementary Table S1.

In Figure 4a, we show a product neural fragility heatmap of the pre and post resection iEEG sessions. The post-resection session is considerably lower in values over the entire network, when compared to the pre-resection session. In this specific patient, Figure 5, shows that E3 had an effect size decrease in neural fragility of 1.43 ± 0.530 (95% CI). The difference between the post and pre resection was significant at *α* = 0.05 with a PValue of 3.12e-12 (K-Sample MANOVA with distance correlation).

**Figure 4.**
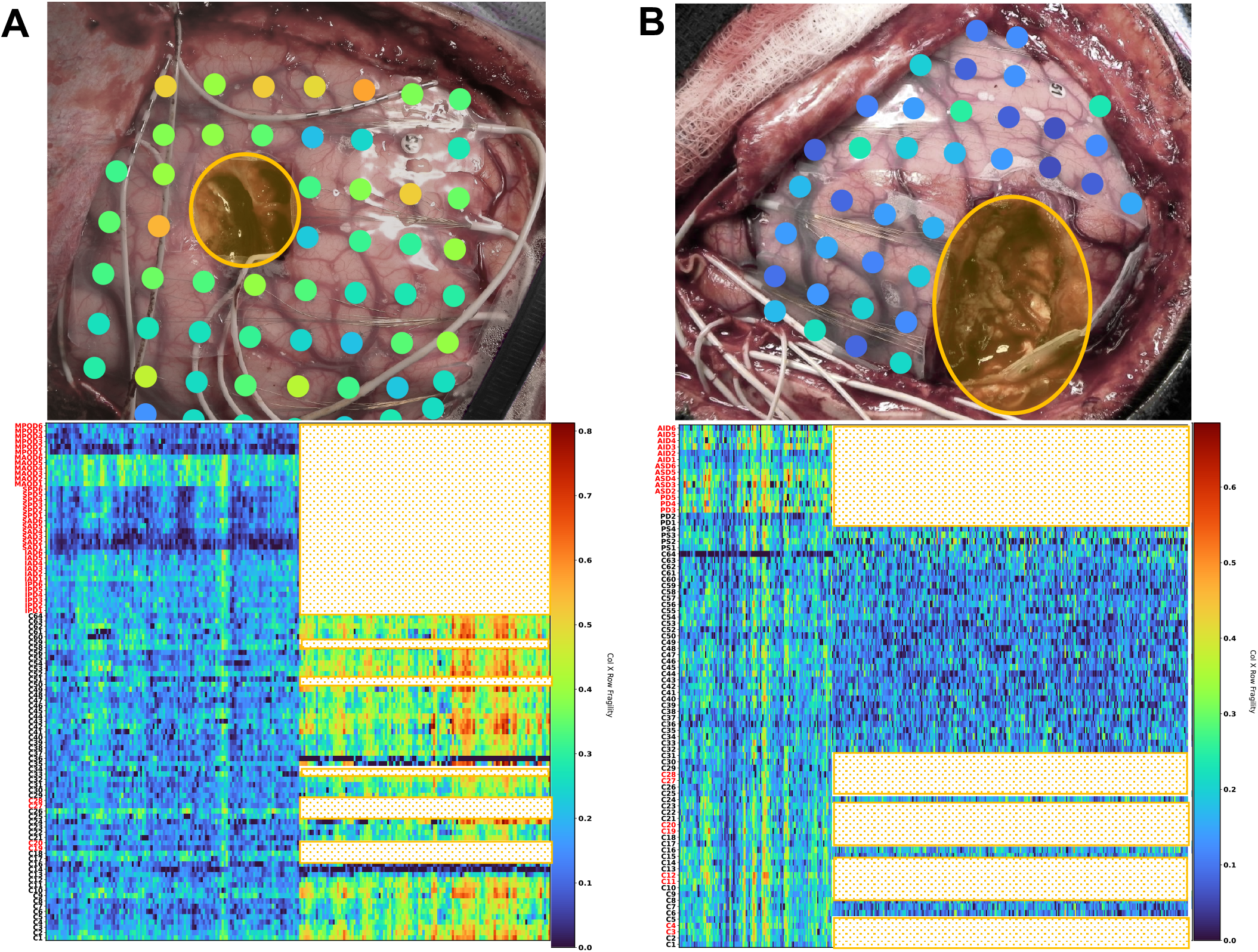
Neural product fragility of successful and failed resections in DRE patients at Sick Children Hospital. **(A)** Resected brain photograph (top) of subject E1 from HSC with Engel III outcome. The heatmap (bottom) shows neural fragility of the pre and post-resection iEEG for a patient with failed resection. The heatmaps show two concatenated sessions: the pre-resection iEEG and post-resection iEEG. Values in the post-resection period go up. **(B)** Resected brain photograph (top) of subject E3 from HSC with Engel I outcome. The heatmap (bottom) shows neural fragility heatmap of the pre and post-resection iEEG for a patient with successful resection. The heatmap shows fragility goes down in the post-resection period. The white region represents the channels that were in the resected regions for the post-resection iEEG simulation, or disconnected due to surgical necessity. The turbo colormap is used in these heatmaps. On the heatmaps’ y-axis, are channel labels, with red channel labels annotated as part of the clinical EZ hypothesis. Note that not all channels are annotated, as some are discarded due to poor recording quality (more information in Supplementary Patient Dataset Collection). In addition, depth electrodes are not visualized as they are all removed as part of the surgical procedure. We analyzed the raw iEEG under a monopolar reference.

**Figure 5.**
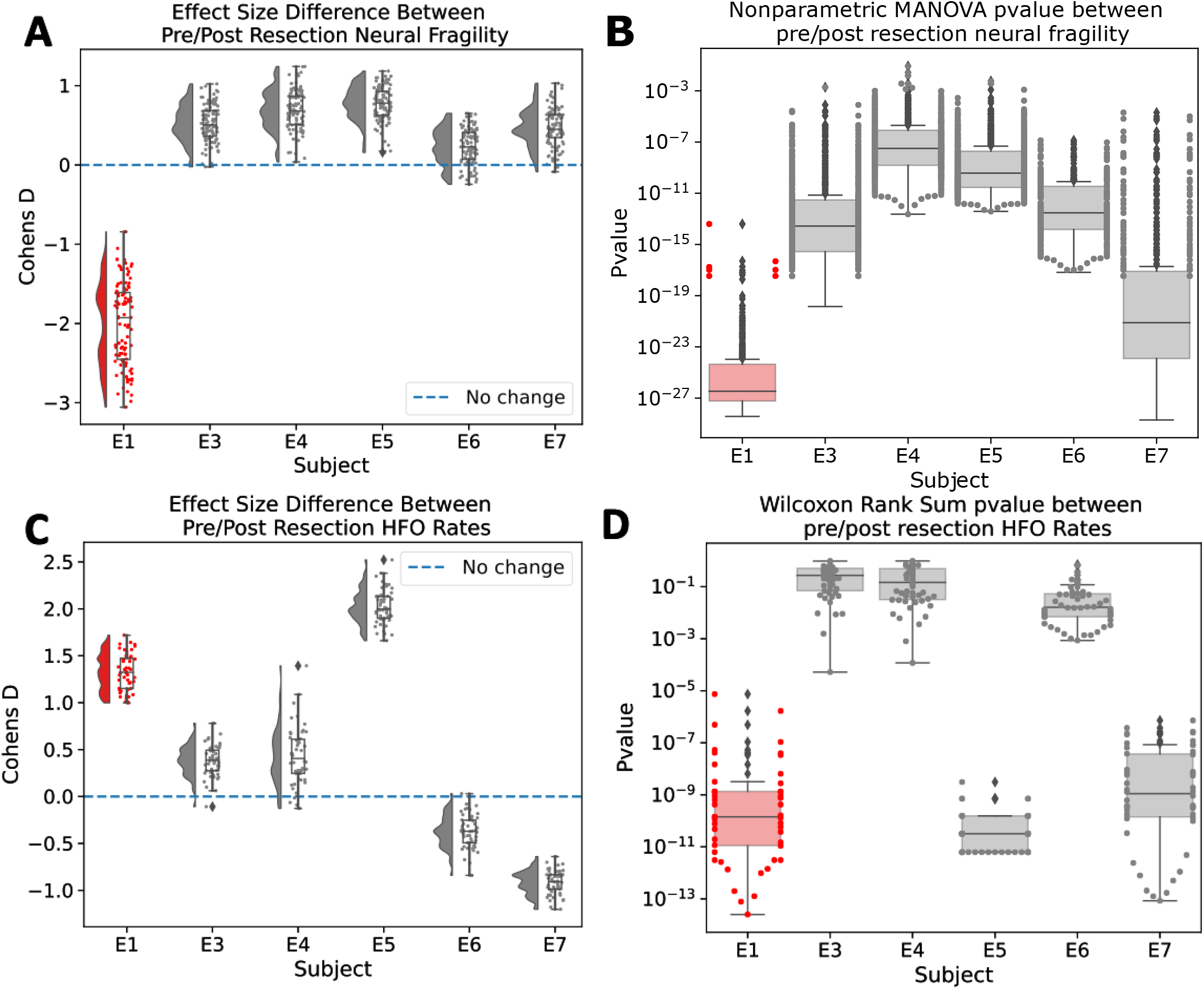
Neural fragility of pre vs post resection effect size differences. **(A)** A summary effect size difference between pre and post resection fragility values for the six patients. Each dot represents the Cohens D effect size computed on a bootstrap sample from pre and post resection heatmap. The successful resections have an improvement in overall network fragility (positive Cohen’s D), while the failed resection shows an actual increase in overall network fragility. **(B)** Showing the distribution of pvalues computed from the same bootstrap samples in (a), that are computed using a K-Sample MANOVA test with alpha level of 0.05. For more information on how the bootstrap procedure was implemented, see Supplementary Statistical Analysis. **(C)** A summary effect size difference between pre and post resection HFO rate values for the six patients. Each dot represents the Cohens D effect size computed on a bootstrap sample from pre and post resection session. A positive effect size indicates that there was a decrease in the HFO rates. HFOs were computed using the RMS detector, described in Supplementary Time Frequency Representation Analysis. **(D)** Corresponding pvalues computed over bootstrap samples of the HFO rates using a Wilcoxon rank-sum test. The graph is displayed on a log-scale on the y-axis.

### 3.4 Neural fragility increases in patient with failed resection

In this dataset, subject E1 had surgical failure with seizure recurrence (Engel III, ILAE 4) after their initial surgical resection. In Figure 4b, we compute the product neural fragility heatmap of the pre and post resection iEEG sessions and report that fragility increases in the post-resection iEEG. From pre to post resection session, the neural fragility of the iEEG network increased by0.567 ± 0.441(mean +/-std of Cohen’s D effect size).

Compared to the other subjects, E1 is the only subject that had an increase in neural fragility after surgery (Figure 5). All other subjects had a decrease in neural fragility ranging from a decrease of 0.845 (subject E7) to 2.324 (subject E5) measured in Cohen’s D effect size. Results were similar if a common average reference was applied to the data as well (see Supplementary Figure S3). All the heatmaps show a marked decrease in neural fragility over the entire network when there was a successful surgical outcome, whereas it increased in the patient with a failed outcome.

### 3.5 Comparing neural fragility and time-frequency spectral features of iEEG

In addition to evaluating neural fragility on the HSC dataset, we also compute common time-frequency based features that would serve as a benchmark. The frequency bands presented and HFOs are common univariate channel features that are looked at by clinicians and the research community in the context of iEEG epilepsy.^21,20,18,12^ See Supplementary Time Frequency Representation Analysis for full details on how we compute these benchmark features. In Figure 5, we compare the pre vs post fragility and compare results to our benchmark features. Fragility is the only one with a clear separation between E1 (i.e. the subject with a failed surgical resection) and patients E3-7 (subjects with Engel I and ILAE 1 surgical outcomes). Moreover, none of the spectral features result in a difference between the pre and post resection sessions (Supplemental Figure S1).

## 4 Discussion

In this study, we analyze neural fragility, a proposed biomarker of epileptogenicity using in-silico and pediatric iEEG. We demonstrate that neural fragility increases/decreases after a failed/successful surgery respectively. We first confirmed this hypothesis in-silico, where we demonstrate how different resection scenarios will affect the overall iEEG network fragility. We then demonstrated the same findings in six pediatric DRE patients from HSC, where all successful surgical patients had a decrease in neural fragility. One patient with a failed surgical outcome showed an increase in overall network fragility.

### Neural fragility compared to traditional proposed features of the EZ

Currently, no prospective definition of the EZ exists.Although HFOs were initially promising,^18,36,37^ the existence of physiological HFOs,^38^ problems with reproducibility of HFO studies^37,36^ and inconclu-siveness of existing clinical trials^12^ suggest that we need to evaluate other approaches. Neural fragility of a neural network approaches the problem of EZ localization from a networked dynamics perspective. Epileptic nodes within a network are hypothesized to cause an imbalance in the network characterized by its network structure. From a biological view, imbalance due to perturbations between excitatory and inhibitory connections of a neural network can occur through any number of mechanisms, such as elevated glutamate,^39^ genetic disorder impacting synaptic inhibition,^40^ decreased GABA,^41^ inclusion of axo-axonic gap junctions,^42^ loss of inhibitory chandelier cells,^43^ or axonal sprouting from layer V excitatory pyramidal cells.^44^ This imbalance within a neuronal network can cause system instability, where impulses at certain nodes lead to recurring seizures. Although iEEG cannot distinguish between excitatory and inhibitory neuronal populations, the concept of imbalance causing the network to be on the brink of instability can be modeled by neural fragility at the iEEG network level. Neural fragility is also resilient to the state of anesthesia, as recordings from HSC were performed intraoperatively under constant total intravenous anesthetic (TIVA).

### Virtual epileptic patients can guide EZ hypotheses

Using TVB, a neuroinformatics platform, we were able to simulate whole-brain activity to pose hypotheses about how a proposed biomarker might modulate as a function of a variety of resections. The advantage of such a platform is that we can simulate data when we know exactly where the EZ is, and then perform in-silico resections by zeroing out the corresponding rows and columns of the structural connectivity matrix. This demonstrates that algorithms, such as neural fragility can be used in conjunction with computational modeling with TVB to explore algorithmic performance in a realistic simulation environment.

### Converging to a prospective definition of the EZ

In this study, we consider the retrospective definition of the EZ: the necessary brain tissue needed to be removed in order to render the patient seizure free. However, other definitions of the EZ have been proposed, such as the epileptogenicity index.^45^ Typically, approaches have tried to either i) build a prediction model for the clinically annotated epileptic channels,^20^ ii) build a prediction model for the clinical annotations on only successful patients^46^ and iii) building a prediction model conditioned on the clinical annotations that predicts surgical outcome.^27^ Building a model to predict the clinical annotations would not obtain a model of the EZ, since clinical annotations are an estimate of the underlying EZ. Such a model would simply replicate clinical behavior at most, which would still result in outcomes between 30-70%.^17^ Building a model using only patients with surgical outcomes would also limit the amount of data one can use. In addition, this approach is limited because clinical annotations are not perfect even in successful outcomes. Finally, building a model conditioned on the clinical annotations to predict surgical outcome takes advantage of both failed and successful outcomes and is a good intermediary. However, it also lacks a direct relationship to the underlying EZ because subject variability can be very high. For example, patients can have seizures recur due to kindling, which would not be necessarily due to the EZ.^47^ Analyzing iEEG data before and after a surgical intervention on the same subject in this study, we have a better estimate of patient-specific EZs because we can observe the post-resection iEEG for seizures and modulation of a proposed biomarker after surgical resection. Combining intraoperative and postoperative monitoring data with large retrospective datasets will be important in evaluating biomarkers for the EZ.

### Outlook of neural fragility and continuous post operative iEEG

Due to the unique nature of the dataset where we have access to iEEG recordings before and after a surgical intervention. As a result, we had the unique opportunity to study how proposed electrophysiological biomarkers change as a result of the surgical intervention. Although our results are encouraging, we have a limited sample size of 6 pediatric patients with only 1 surgical failure. Future studies should expand on these findings, particularly as they pertain to surgical failures to validate the proposed biomarker. In Li et al.,^27^ neural fragility was validated on 91 adults (18 years or older). However, these patients were all children under the age of 18, and thus future research would have to further validate that these results hold for adults.

The paradigm of intraoperative continuous iEEG monitoring to obtain pre and post-resection iEEG data of the same set of electrodes presents an opportunity to study dynamics as a result of surgical interventions. However, clinical epilepsy is moving towards stereotactic EEG (sEEG) implantations and the surgical procedure for keeping electrodes implanted during a resection has not been developed. If sEEG data can be obtained, caution should be taken to annotate white matter contacts.48 Future clinical developments that enable similar pre and post-resection sEEG data will be important to see if neural fragility still modulates with respect to surgical outcomes.

## Data Availability

Due to the unique nature of the data from the hospital for Sick Children and HIPAA concerns, the dataset cannot be made publicly available. Instead the data is available upon request from the clinical co-authors. In addition, the TVB dataset used in simulations is accessible upon request from Marseille University.

## 6 Acknowledgements

AL is supported by NIH T32 EB003383, the NSF GRFP (DGE-1746891), the ARCS Scholarship, Whitaker Fellowship and the Chateaubriand Fellowship. SVS is supported by NIH R21 NS103113, Maryland Innovation Initiative and the Burroughs Wellcome Fund CASI Award 1007274. VJ has received funding from the European Union’s Horizon 2020 Framework Programme for Research and Innovation under the Specific Grant Agreement No. 945539 (Human Brain Project SGA3). The authors would like to thank Macauley Breault for help in creating brain figures.

## 7 Author Contributions

AL, GI, VJ and SVS conceived the project. NW and GI oversaw data collection from The Hospital for Sick Children. VJ provided TVB data from Marseille University and guidance on TVB simulations. AL organized the data, converted to BIDS, conducted the analyses and generated the figures. AO, HO annotated the data and marked clips of data to be used. KG, NW provided input on the data and experimental design. SK localized electrode coordinates and performed co-registration to obtain brain figures. PM helped run HFO experiments. AL wrote the paper with input from the other authors.

## 10 Code availability

Code used for reproducing the figures and running parts of the analysis are at https://github.com/adam2392/sickkids.

## Supplementary Materials and Methods

### 10.1 Patient Dataset Collection

#### 10.1.1 The Hospital for Sick Children

iEEG data from 7 DRE patients were analyzed. These patients underwent intracranial EEG monitoring between January 2017 and December 2019 at The Hospital for Sick Kids (HSC). All children underwent subdural grid implantation and the same chronically implanted electrodes were used without changing position or orientation for extraoperative and subsequent intraoperative electrocorticography, using a previously reported technique.^1^

That is, the subdural electrodes were placed in the first stage procedure and extraoperative mapping took place over the span of a one-week hospital admission. The patients then underwent a second stage (resective) procedure while the same electrodes in the identical position continued to record in real-time while the resection took place. One patient (E2) was excluded from analysis due to technical challenges during the surgery with the post-resection iEEG. Data were recorded using a Natus (Pleasanton, CA) acquisition system with a sampling rate of 2048 Hz.

Signals were referenced to a common electrode placed subcutaneously on the scalp, on the mastoid process, or on the subdural grid. In ictal recordings, the time of seizure onset was indicated by a variety of stereotypical electrographic features which included, but were not limited to, the onset of fast rhythmic activity, an isolated spike or spike-and-wave complex followed by rhythmic activity, or an electrodecremental response. We discarded electrodes from further analysis if they were deemed excessively noisy by clinicians, recording from white matter, or were not EEG related (e.g. reference, EKG, or not attached to the brain). All patients underwent neuropsychological assessment prior to invasive monitoring, which included measures of Full-Scale intelligence quotient (IQ), verbal comprehension, visual spatial reasoning, visual fluid reasoning, working memory, and visual processing speed using the Wechsler Intelligence Scale for Children (WISC-V) and the corresponding WISC-V sub-tests. Verbal memory was indexed by a child’s overall performance in delayed free recall using the Children’s Memory Scale (CMS), Children’s Auditory Verbal Learning Test-2 (CAVLT-2), or the Child and Adolescent Memory Profile (ChAMP).^2^ Visual memory was indexed by the delayed free recall using the CMS. The FSIQ scores for each patient can be found in Supplementary Table. Detailed neuropsychological profiling may be found in Supplementary Table.

We define successful outcomes as seizure free (Engel class I and ILAE scores of 1 and 2) at 12+ months post-op and failure outcomes with seizure recurrence (Engel classes 2-4).

#### 10.1.2 The Virtual Epileptic Patient

SEEG electrodes were implanted in the regions suspected to be in the EZ. Each electrode had 10–15 contacts (length: 2 mm, diameter: 0.8 mm, contacts separation: 1.5 mm). To determine electrode positions, an MRI was performed after electrodes implantation (T1 weighted anatomical images, MPRAGE sequence, TR = 1900 ms, TE = 2.19 ms, 1.0 × 1.0 × 1.0 *mm*^3^, 208 slices) using a Siemens Magnetom Verio 3T MR-scanner. To reconstruct patient specific connectomes (DTI-MR sequence, angular gradient set of 64 directions, TR = 10.7 s, TE = 95 ms, 2.0 × 2.0 × 2.0 *mm*^3^, 70 slices, b weighting of 1000 s/*mm*^2^, diffusion MRI images were also obtained on the same scanner. The study was approved by the Comité de Protection (CPP) Marseille 2, and all patients signed an informed consent form.

To quantify the proximity and number of tracks between electrodes, structural and diffusion MRI data were obtained via a processing pipeline to derive individualized cortical surface and large-scale connectivity. Cortical and subcortical surfaces were reconstructed along with volumetric parcellations using the Desikan–Killiany atlas, with the cortical regions subdivided in four (280 cortical regions and 17 subcortical regions). We obtained electrode positions by coregistering the parcellation with the MRI scan, and assigning each contact to the region containing most of the reconstructed contact volume. To compute the number of tracks between electrodes, head-motions and eddy-currents were corrected in diffusion data. Fiber orientation was estimated with constrained spherical deconvolution, and 2.5×106 streamlines were obtained by probabilistic tractography. We used the anatomically-constrained tractography (ACT) and the spherical-deconvolution informed filtering of tractograms (SIFT) frameworks to improve reproducibility and biological accuracy. The number of tracks between two pairwise electrodes was then obtained by summing the number of tracks whose extremities belong to the corresponding pairwise brain regions.

### 10.2 iEEG Data Preprocessing

Data was initially stored in the form of the European Data Format (EDF) files.^3^ We preprocessed data into the BIDS-iEEG format and performed processing using Python3.6, Numpy, Scipy, MNE-Python and MNE-BIDS.^4,5,6,7,8,9,10^

Every dataset was notch filtered at 60 Hz and its corresponding harmonics (with a cutoff window of 2 Hz), and bandpass filtered between 0.5 and the Nyquist frequency with a fourth order Butterworth filter. If correlated noise was present, a common average reference was applied. EEG sequences were broken down into sequential windows and the features were computed within each window (see Neural Fragility Analysis for details). Values at each window of time were normalized across electrodes to values that could range from 0 up to at most 1, to allow for comparison of relative feature value differences across electrodes over time; the higher a normalized feature, the more we hypothesized that electrode was part of the EZ.^11^ This normalization scheme allows us to account for how relatively different channels are in terms of the proposed metric relative to other channels over time.

Channels with significant artifact were excluded. Artifact-free data segments of equal length were selected pre and post resection of the epileptogenic zone. As part of the surgical resection, certain channels were disconnected to allow the surgeons to get at certain tissue. These channels are represented as “NaN” recordings. Moreover, since surgical resection occurs, post-resection recordings will have then less channels compared to their pre-resection counterparts.

### 10.3 Neural Fragility Analysis

To compute fragility heatmaps from iEEG recordings, we first constructed simple linear time-invariant models in a small window of time. This is analogous to a simple, linear dynamic causal model.12 We partition the iEEG data into 250 millisecond windows, and estimate a linear time-invariant model, giving us an A matrix. This A matrix represents how dynamics of the iEEG evolve in that small window. For each A matrix, we compute the minimum perturbation necessary to apply to each channel’s connections to destabilize the system, i.e., move the eigenvalues outside the stable region. We repeat this for every channel, which gives us a neural fragility values for all channels over all the time windows.

To compute fragility heatmaps from iEEG recordings, we first constructed simple linear models as described above and in equation 1.

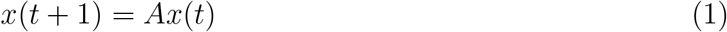

Since each observation, *x* ∈ ℝ^*d*^, has dimension d (number of channels), we would like to formulate a least-squares estimation procedure with *n* > *d* samples. We choose n to represent a 250 ms iEEG window. We then have the following representation of *X*(*t*) ∈ ℝ^*d*×*n*−1^:

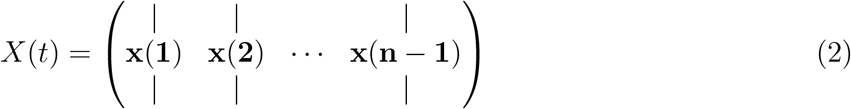

and the following representation for *X*(*t* + 1) ∈ ℝ^*d*×*n*−1^

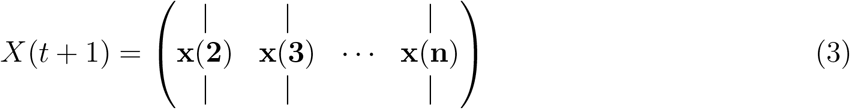

The least-squares will now seek to fit a linear operator *A* such that:

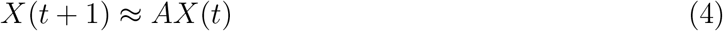

This linear operator representation of the dynamical system has connections to Koopman operator theory^13^ and Dynamic Mode Decomposition in Fluid Mechanics.^14^ We seek to approximate the inherently nonlinear iEEG dynamics within a small window of time using a finite-dimensional approximation of the Koopman operator using the observables (i.e. *x*(*t*)) themselves. We specifically used least-squares algorithm with a 10e-5 l2-norm regularization to ensure that the model identified was stable (with absolute value of eigenvalues ≤ 1) as in.^15,11^ Then, we slid the window 125 ms and repeated the process over the entire data sample, generating a sequence of linear network models in time.

We systematically computed the minimum perturbation required for each electrode’s connections to produce instability for the entire network as described in.^11^ This is represented in equation 5, where the Δ_*i*_ is the desired column perturbation matrix for channel i, and the *λ* = *r* ∈ ℂ is the desired radii to perturb one single eigenvalue to.

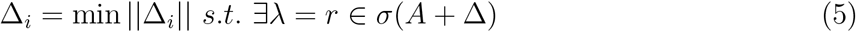

More specifically, we compute a structured perturbation matrix, such that:

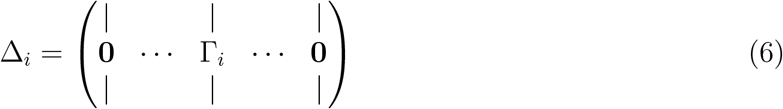

where each Γ_*i*_ ∈ ℝ_*d*_ is the actual column perturbation vector. The intuition for using this type of structured perturbation is described in the main paper in Neural fragility of an iEEG network. We demonstrate how to solve for this using least-squares in.^11^ The electrodes that were the most fragile were hypothesized to be related to the EZ in these epilepsy networks (seen as the dark-red color in the turbo colormap in Figure 4).

### 10.4 Time Frequency Representation Analysis

We computed HFOs using a variety of methods. HFOs were computed using https://github.com/adam2392/hfo, an open-source Python implementation of HFO detection algorithms.16 The Line Length, RMS, Hilbert and Morphology detectors are used.^17,18,19^ We defined HFOs as the union of ripples (80-250 Hz) and fast ripples (250-500 Hz) as detected by the RMS detector.

We also constructed frequency-based features from frequency bands of interest by applying a multi-taper Fourier transform over sliding windows of data with a window/step size of 2.5/0.5 seconds.^20^ We required relatively longer time windows to accurately estimate some of the lower frequency bands. Each EEG time series was first transformed into a 3-dimensional array (electrodes X frequency X time), and then averaged within each frequency band to form six different spectral feature representations of the data. We break down frequency analysis as follows:

1. Delta Frequency Band [0.5 - 4 Hz]
2. Theta Frequency Band [4 - 8 Hz]
3. Alpha Frequency Band [8 - 13 Hz]
4. Beta Frequency Band [13 - 30 Hz]
5. Gamma Frequency Band [30 - 90 Hz]
6. High-Gamma Frequency Band [90 - 300 Hz]
7. HFO = R & FR [80-250 Hz & 250-500 Hz]

### 10.5 The Virtual Brain Patient-Specific Modeling

To simulate a resection of a region, we removed the connections of that region to all other regions and then generated the resulting iEEG data.

We utilize the Epileptor model, where it was originally designed to produce realistic seizure dynamics.^21^ We specifically, use an extension of the original Epileptor model, called the resting-state Epileptor, which is capable of reproducing interictal spikes.^22,23,24,25,26^ The resting-state Epileptor equations are as follows:

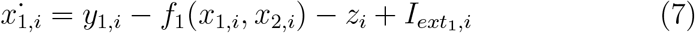

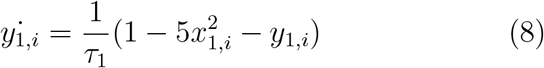

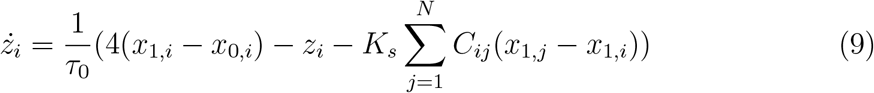

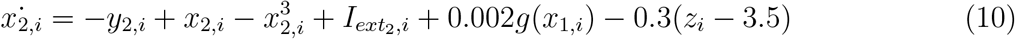

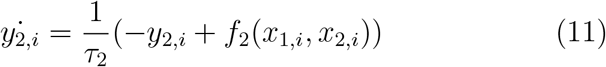

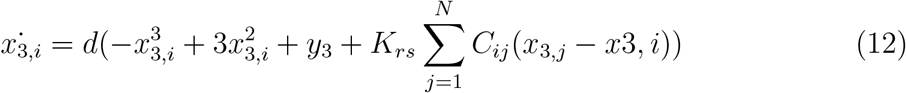

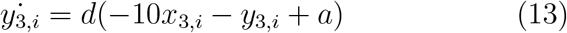

where

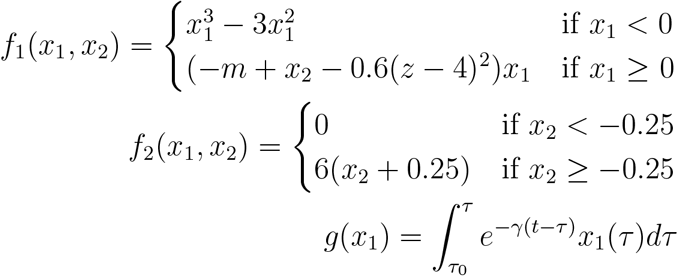

The output that is measured in the original Epileptor model is the LFP, defined as *x*_2_ − *x*_1_. In the resting-state Epileptor model, the output is defined as a convex combination of the fast and intermediate subpopulation activity and the resting-state subpopulation activity.

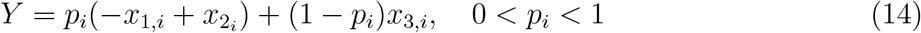

The *i* indexes the *N* discrete brain regions (i.e. 84 brain regions in a Desikan-Killiany atlas). Here, the *x*_1_, *y*_1_ variables correspond to the fastest time scale accounting for low-voltage fast discharges (i.e. very fast oscillations). The *x*_2_, *y*_2_ variables correspond to an intermediate time scale accounting for spike-and-wave discharges. The *z* slow-permittivity variable corresponds to the slowest time scale, responsible for autonomously switching between interictal and ictal states in the form of a direct current (DC) shift.^27,28,21^ This variable takes the system through saddle-node and homoclinic bifurcations for seizure onset and offset respectively. The *x*_3_, *y*_3_ variables account for transient behavior, in the form of spindle-like patterns, which added allow the Epileptor model to reproduce resting-state oscillatory wave patterns and also reproduce interictal spikes. The *x*_0_ serves as a hyperparameter, denoting the degree of epileptogenicity of a brain region. If *x*_0_ is greater than a critical value, of -2.05, then the brain region can trigger seizures autonomously. Otherwise, it is in an equilibrium state. The *a* hyperparameter relative to the critical value of -1.74, also represents the degree of epileptogenicity during the interictal resting state. The *C*_*ij*_ are the weights based on the subject’s structural connectivity matrix and *K*_*s*_, *K*_*rs*_ are the respective large-scale scaling parameters of the connecitivity weights in the seizure and resting-state subpopulations. Note that *C*_*ij*_ = 0, ∀*i* = *j* because we assume that the neural mass model of one brain region already accounts for internal connectivity effects. The interictal and preictal spikes occur when these variables are excited by the fast oscillation system via the coupling term, *g*(*x*_1_). The characteristic frequency rate d, fixed to 0.02, sets the natural frequency of the third subsystem (10 Hz), the most powerful frequency peak observed in electrographic recordings at rest.^29,30^ For more detailed discussion on the Epileptor and extensions, see.^21,22,31^

In our system, we set a range of *x*_0_ value combinations for the EZ and propagation regions, with *x*_0_ = −2.10 for the clinically hypothesized EZ region. Then *x*_0_ = −2.35 was set for the normal region for all simulations. Our goal here is to match the real data situation as closely as possible, where we are not comparing seizures, but resting-state iEEG activity. The *a* variable is set to -1.74. Finally, *p*_*i*_ = 0.2. We set EZ regions based on what the clinicians thought for this patient. The other parameters, *τ*_0_ = 4000; *τ*_2_ = 10; *K*_*s*_ = −5; *γ* = 0.01 and then the rest of the parameters were set as in.^26^

The system of stochastic differential equations were solved using an Heun Stochastic integration scheme with an integration step of 0.05, which gave values of the local field potentials in 14. To start the simulation at a realistic point, initial conditions were computed and stored for each simulation using a burn-in period of 15 seconds. Simulations were 30 seconds long afterwards. Additive white Gaussian noise was introduced into the variables *x*_2_ and *y*_2_ with mean 0 and variance of 0.0005 and to *x*_3_ with a mean of 0 and variance of 0.0001. Additionally, observational colored noise was added to the iEEG simulated data with a mean of 0 variance of 1.0 and noise correlation (i.e. color) in time of 0.1. The iEEG data was modeled using a forward solution that uses an inverse gain mean-field model from the LFP. The details of which are described in.^26^

To simulate a “virtual” resection, we took the actual clinical resection performed, and “removed” those regions in the structural connectivity matrix for that patient. This corresponded to “zeroing” out those rows and columns for that brain region, simulating the removal of that region. When we remove these brain regions, we also virtually removed the corresponding iEEG channels in those brain regions. We then used the above setup to simulate neural dynamics and corresponding iEEG activity pre and post resection. Afterwards, these snapshots of data can be analyzed using neural fragility as described in Neural Fragility Analysis. They are then compared as described in Statistical Analysis.

### 10.6 Statistical Analysis

To compare pre and post resection data, we computed Cohen’s D effect size differences between the feature representations of these two data sessions. We use the Mann-Whitney U test and K-Sample MANOVA test to compute a PValue with *α* Type I error rate set to 0.05. The distance function utilized in the K-Sample MANOVA is the distance correlation function, which is a more robust version of Pearson correlation. The null hypothesis of our experimental setup was that the pre resection metrics came from the same population as the post resection. The alternative hypothesis was that the populations were different.

Since the feature representation heatmaps (i.e. neural fragility spatiotemporal heatmap) show a metric over time of all the recorded channels, samples are not necessarily independent. To account for correlations in time, we use a contiguous bootstrapping procedure to estimate the Cohen’s D and PValues with n=100 bootstrap samples. That is the typical bootstrap algorithm is carried out along the time-axis. Each bootstrap sample in time consists of all the channels, along with a small window of 10 samples (i.e. about 1 second). Then these bootstrap samples are compared between the pre and post resection heatmaps, generating a bootstrap distribution with a reported mean and standard deviation with 95% confidence intervals typically reported.

## Tables

**Table S1:** A table of patients in study from the Hospital for Sick Children. There was a subject E2 with similar types of procedure done, but the dataset had different electrodes implanted before and after resection, rather then the same electrodes as with the rest of patients. We categorized patients by their clinical complexity (CC) as follows: (1) lesional, (2) focal temporal, (3) focal extratemporal, and (4) multifocal.^32,33^ If a patient presents with multiple categories, they are filed in the more complex case. Some patients presented with focal cortical dysplasia (FCD).

| Patient Summary |  |  |  |  |  |  |  |  |  |  |
| --- | --- | --- | --- | --- | --- | --- | --- | --- | --- | --- |
| Subject | Pathology | Clinical complexity | Engel | ILAE | Ethnicity | Age | Onset Age | Handedness | Sex | Months Follow Up |
| E1 | FCD type IIb | 4 | III | 4 | Caucasian | 6-10 | 1-5 | R | M | 31 |
| E3 | FCD type IIb | 1 | I | 4 | Asian | 11-15 | 6-10 | R | F | 13 |
| E4 | FCD type IIb | 1 | I | 4 | Caucasian | 11-15 | 11-15 | L | M | 21 |
| E5 | FCD type IIa and IIb | 4 | I | 4 | Asian | 11-15 | 6-10 | R | M | 31 |
| E6 | FCD type IIb | 1 | I | 4 | Caucasian | 11-15 | 6-10 | R | F | 14 |
| E7 | left amygdala enlargement and hippocampal signal changes | 2 | I | 4 | LatinX | 16-20 | 11-15 | R | M | 10 |

**Table S2:** A table of patients in study used for TVB simulations.

| Patient Summary |  |  |  |  |  |  |  |
| --- | --- | --- | --- | --- | --- | --- | --- |
| Subject | Clinical complexity | Engel | Age | Onset Age | Handedness | Sex | Months Follow Up |
| TVB001 | 2 | I | 36-40 | 6-10 | n/a | M | 12 |

## Supplementary Figures

**Figure S1:**
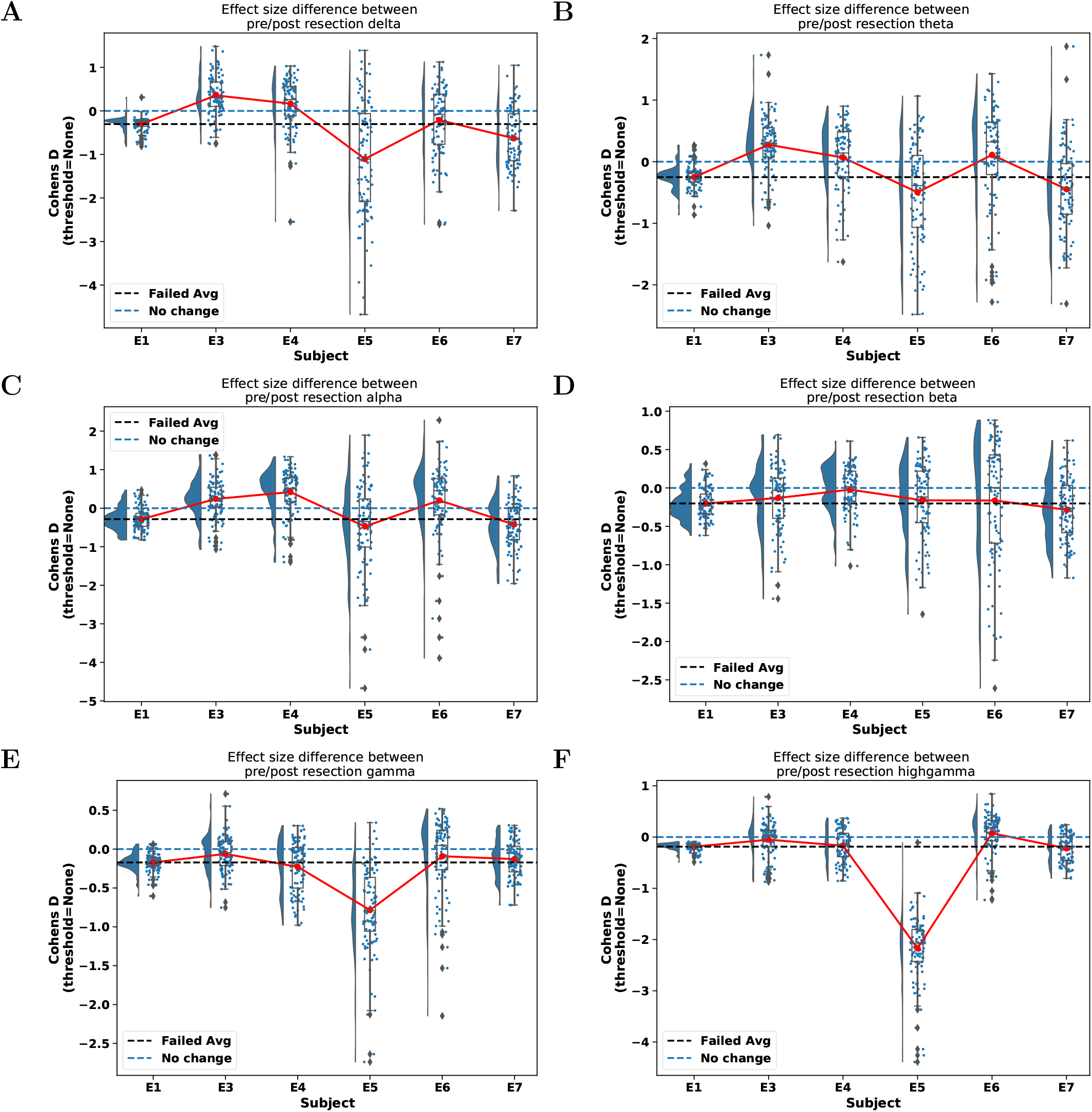
Pre vs post resection effect size plots of power in frequency bands. **(a)-(f)** Are delta, theta, alpha, beta, gamma, and highgamma frequency bands respectively. For full details on computing the frequency band power heatmaps, see Time Frequency Representation Analysis. For examples of the time-frequency heatmaps for all subjects and all frequency bands computed, see Supplementary files.

**Figure S2:**
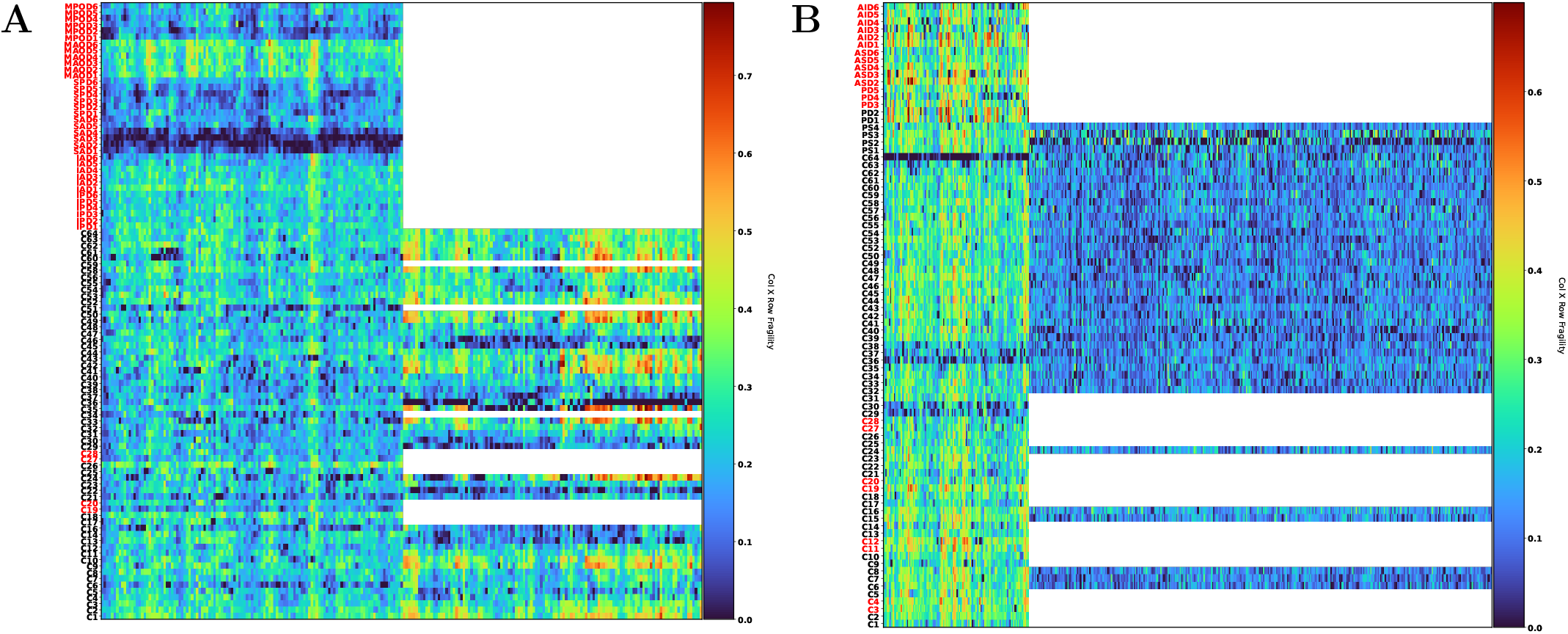
Product neural fragility heatmaps using common average referencing. **(A)** Subject E1 with common average reference and **(B)** subject E3 with common average reference. This is the same heatmaps over the same period of data as Figure 4, but using a different reference on the data.

**Figure S3:**
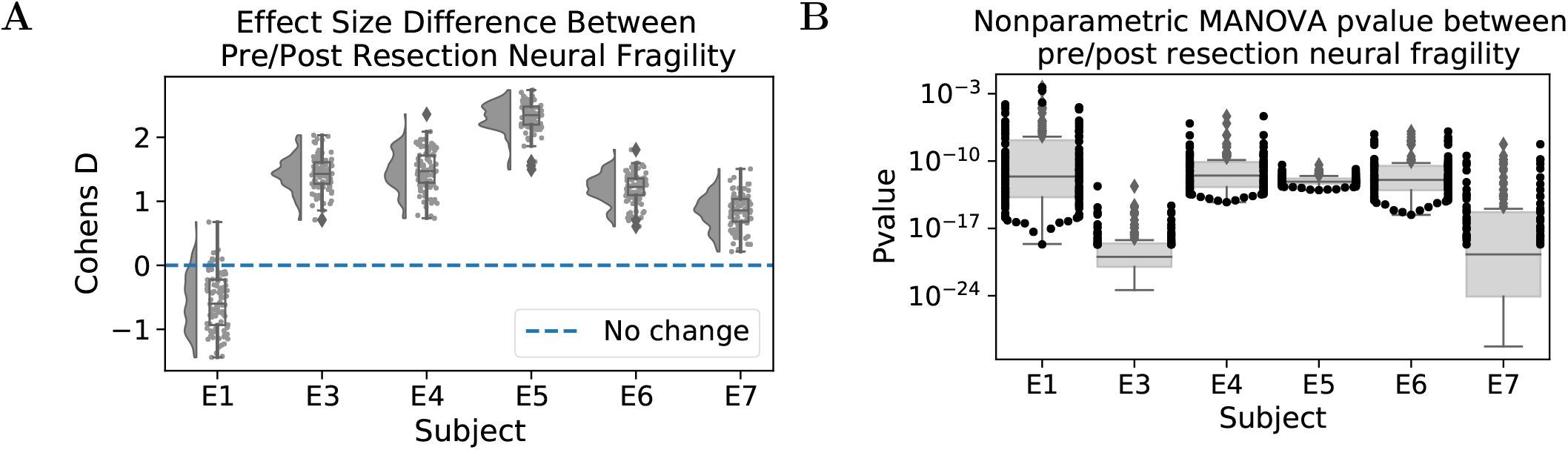
Cohen’s D effect size and p-value comparisons of product fragility with common average reference. **(A)** Cohen’s D effect size comparisons between pre and post resection product neural fragility. **(B)** Non-parametric KSample p-values computed between pre and post resection product neural fragility. This is the same period of data and procedure as Figure 5, but with a common average reference applied to the data.

